# Strategies to deliver smoking cessation interventions during targeted lung health screening - a systematic review and meta-analysis

**DOI:** 10.1101/2023.03.28.23287843

**Authors:** Parris J Williams, Keir EJ Philip, Saeed M Alghamdi, Alexis M Perkins, Sara C Buttery, Michael I Polkey, Anthony A Laverty, Nicholas S Hopkinson

**Author notes:** Professor Nicholas S. Hopkinson (Corresponding author) National Heart and Lung Institute, Imperial College, Royal Brompton Hospital Campus, Fulham Road, London SW3 6HP.

## Abstract

**Introduction:** Lung cancer screening presents an important teachable moment to promote smoking cessation, but the most effective strategy to deliver support in this context remains to be established.

**Methods:** We undertook a systematic review and meta-analysis of smoking cessation interventions delivered during lung health screening, published prior to 20/07/2022 MEDLINE, PsychINFO, CENTRAL, EMBASE, CINAHL and Scopus databases. Two reviewers screened titles, and abstracts, four reviewed each full text using prespecified criteria, extracted relevant data, assessed risk of bias and confidence in findings using the GRADE criteria. The review was registered prospectively on PROSPERO (CRD42021242431).

**Results:** 10 randomised controlled trials (RCT) and 3 observational studies with a control group were identified. Meta-analysis of 9 RCTs demonstrated that smoking cessation interventions delivered during lung screening programmes increased quit rates compared to usual care (OR: 2.01, 95%: 1.49-2.72 p<0.001). 6 RCTs using intensive (≥3 behavioural counselling sessions) interventions demonstrated greater quit rates compared to usual care (OR: 2.11, 95% CI 1.53-2.90, p<0.001). A meta-analysis of 2 RCTs found intensive interventions were more effective than non-intensive (OR: 2.07, 95%CI 1.26-3.40 p=0.004), Meta-analysis of 2 RCTs of non-intensive interventions (≤2 behavioural counselling sessions or limited to online information audio take home materials such as pamphlets) did not show a higher quit rate than usual care (OR: 0.90, 95% CI 0.39-2.08 p=0.80).

**Discussion:** Moderate quality evidence supports smoking cessation interventions delivered within a lung screening setting compared to usual care, with high-quality evidence that more intensive interventions are likely to be most effective.

## INTRODUCTION

Tobacco use remains a primary driver of global morbidity, mortality, and related economic and healthcare costs, responsible for more than 8 million deaths per year(1, 2). Despite mortality rates declining for most cancers, lung cancer deaths have remained high, accounting for 21% of all cancer deaths in the UK (3). Patients typically present to healthcare services at a late stage, resulting in late diagnosis and a poorer prognosis(3). Early diagnosis through lung cancer screening programmes has achieved success in the USA and Europe. For example, the National Lung Cancer Screening trial in the US saw a 20% reduction in lung cancer mortality among their populations who underwent screening(4). In 2019 the NHS rolled out their Targeted Lung Health Check (TLHC) pilot project with 77% of lung cancers identified being stage 1 and 2 (5) and lung cancer screening has recently been recommended by the UK National Screening Committee(6).

Eligibility criteria for lung screening programmes, vary slightly between countries, however participants are typically: middle aged (55–75), a current or former smoker (quitting within the last 15 years), with a 30+ pack year history. Around 40-50% of people taking part still smoke(7), and are likely to be highly nicotine dependent creating a significant barrier for cessation(8). Attendance at screening programmes has been highlighted as a teachable moment for smoking cessation, with several randomised controlled trials (RCTs) of screening demonstrating increased quit rates among individuals taking part (8–11), however there is substantial scope to increase the quit rates observed (11-15%).

Increasing quit rates among this high-risk population has the potential to dramatically increase the impact and cost effectiveness of screening programmes, beyond their effect on early identification of lung cancers, by reducing the risk of future smoking related illness(12, 13). Recent modelling estimates that 1.1 million smokers, around 15.4% of all the people in England who smoke, will meet the criteria for screening, representing a potentially huge public health impact (14). Identifying the most effective strategies for delivering cessation within a lung screening context is now recognised as a priority by multiple health organisations including the American Thoracic Society, NHS England, and the European Respiratory Society (15–17). Studies investigating the effects of delivering smoking cessation interventions in this context vary in terms of intervention type, frequency, and intensity of intervention. A systematic review published in 2016 concluded that lung screening programmes were optimal environments to deliver smoking cessation, however did not offer evidence on what type of smoking cessation delivery modality is most effective (11).

This area of screening and smoking cessation medicine is rapidly evolving, and despite organisations across Europe and the US recommending that cessation support be incorporated into lung screening programmes, they do not provide exact guidelines on what form this should take. We therefore conducted a systematic review and meta-analysis to determine the most effective delivery strategies for smoking cessation interventions conducted during lung health screening programmes.

## METHODS

The Preferred Reporting Items for Systematic Reviews and Meta Analyses (PRISMA) guidelines were used to complete and report this systematic review, which was registered prospectively on PROSPERO(CRD42021242431).

### Inclusion criteria

1. Study type: randomized controlled trial (RCTs) and observational studies with a control group.
2. Population: current smokers enrolled in a lung health screening program, including any of the following interventions: Low Dose Computed Tomography (LDCT) scanning, thoracic radiograph, spirometry assessments, formal clinic visit with a physician specialising in respiratory health.
3. Intervention: a smoking cessation program (e-cigarette, behavioural therapy, pharmacological, motivational interviewing, group counselling, telephone counselling, online materials, and information leaflets).
4. Control: usual care or comparator as defined by the study.
5. Outcome measures: smoking quit rates, quit attempts and reductions in cigarettes smoked.

### Exclusion criteria

1. Studies not published or translated into English.
2. Abstracts, conference posters, expert opinion, review articles, study protocols and student dissertations.

### Search Strategy

We searched MEDLINE, PsychINFO, CENTRAL, EMBASE, CINAHL and Scopus from 1950- 20^th^ July 2022. Our search strategy used exploded medical subject headings in the following: smoking OR tobacco OR vape or vaping OR e-cigarette OR cigar OR nicotine AND cessation OR cease OR stop OR quit OR intervention OR abstain OR abstinence OR relapse OR reduce OR give up AND screen OR scan OR check OR early detection OR early diagnosis OR CT) ADJACENCY2 lung OR thoracic OR pulmonary AND early detection of cancer OR tomography, emission- computed OR mass screening OR spirometry. We also reviewed reference lists of included studies.

### Data extraction and quality assessment

Two researchers (PW & NSH) reviewed all titles and abstracts for eligibility. Four independent reviewers (PW, KEJP, SA, AP) read full text studies to decide on final inclusion and exclusion (Figure 1). Discrepancies were resolved by discussion between all co-authors. Data were extracted by PW and AL using a predefined standardised form and COVIDENCE software. Data recorded included study design and setting, methodology, participant baseline characteristics, type and intensity of smoking cessation intervention used, definition of control group, primary and secondary endpoints, measurement of abstinence and follow up period. Quality of the evidence including the risk of bias (ROB) was measured by two independent reviewers (PW and AL) using the GRADE criteria (complete list of extracted data and GRADE criteria appraisal provided in online supplement)(18). Studies were grouped by outcome assessment and intensity of the intervention. We defined intensive sessions as ≥3 behavioural counselling sessions and non-intensive interventions as those consisting of ≤2 behavioural counselling sessions or limited to online information audio take home materials such as pamphlets.

**Figure 1.**
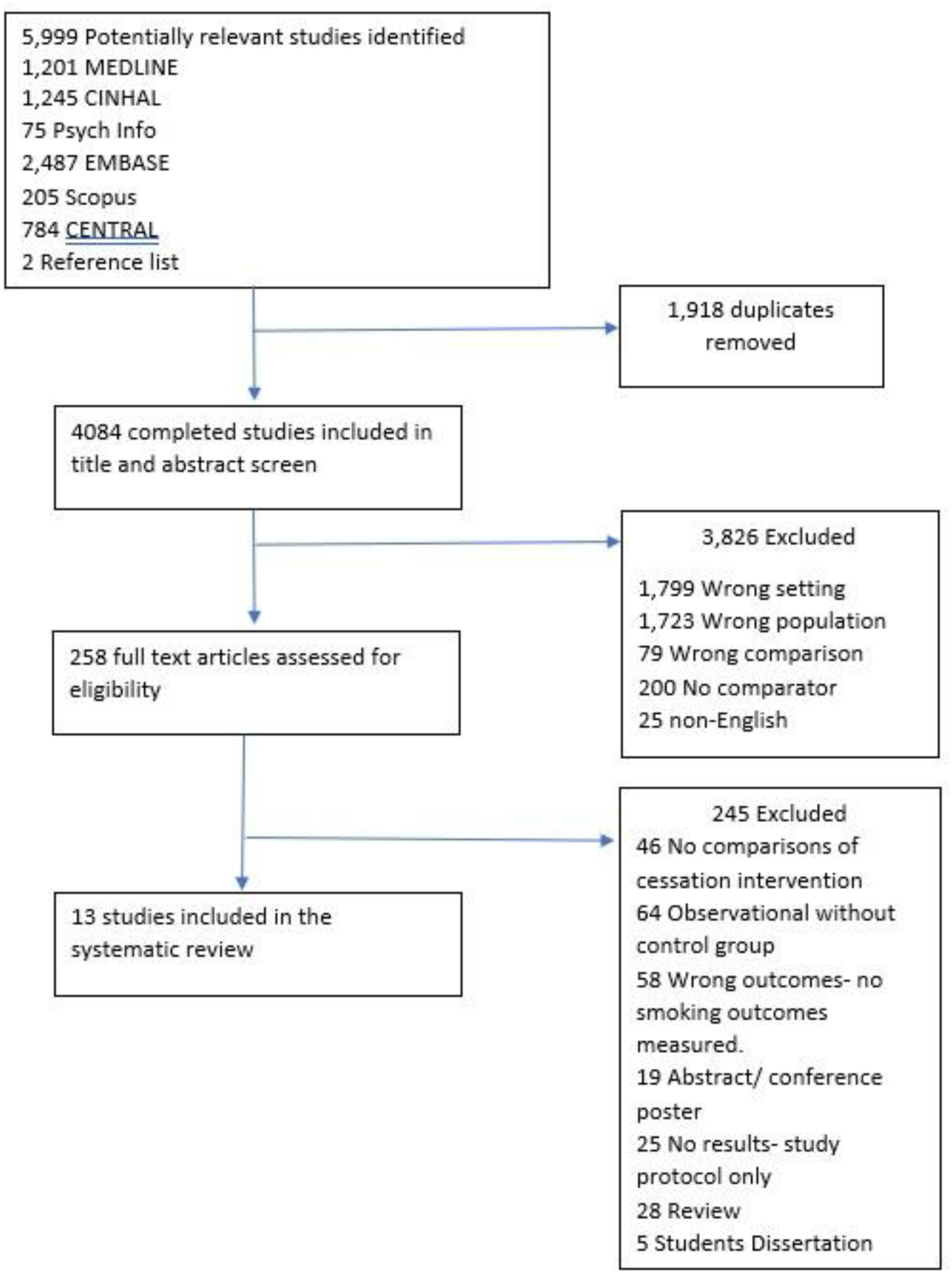
Prisma, study flow diagram.

### Outcome measures Primary outcomes

We considered smoking abstinence at the end of the trial study period (acknowledging that this could vary from trial to trial) as our primary outcome. Smoking abstinence could be defined as; the self-reporting of abstinence of smoking cigarettes in the past 7 days or more, following the cessation intervention, with or without biochemical verification. The measure of abstinence could include, point prevalence, prolonged or continuous and biochemical verification could include exhaled carbon monoxide (CO) or urine cotinine. All these abstinence measures are recommended by Piper et al’s., 2020 review for defining and measuring smoking abstinence in clinical trials(19). To evaluate the effect of interventional intensity, we compared quit rate among studies that provided intensive interventions and non-intensive interventions, compared with control. We also assessed quit rates among studies that provided pharmacotherapy (NRT, E-cigarettes or prescription medications).

### Secondary outcomes

Secondary outcomes included changes in smoking behaviour in terms of number of quit attempts and reductions in tobacco usage (daily cigarette consumption). We defined quit attempts as the number of individuals self-reporting a quit attempt post cessation intervention and reduction in tobacco use as a reduction in the number of self-reported daily cigarettes from baseline.

We searched for information on intervention impacts in terms of long-term mortality, exacerbations of respiratory diseases and patient experience, however none of the screened or included articles measured these as endpoints.

### Data Analysis

A fixed effects meta-analysis was performed to estimate the pooled differences and 95% CI in quit rate and quit attempts between smoking cessation intervention groups and control group. A Mantel-Haenszel fixed effects model was used to obtain estimates for dichotomous data, expressed as odds ratios (OR). To answer our primary and secondary questions we performed the following meta-analyses on data from included RCT; quit rate between smoking cessation interventions vs usual care, quit rate between intensive smoking cessation interventions vs usual care, quit rate between intensive smoking cessation interventions + pharmacotherapy vs usual care, quit rate between non-intensive smoking cessation interventions vs usual care, quit rate between intensive + pharmacotherapy smoking cessation interventions vs non-intensive interventions, and quit attempts between smoking cessation interventions vs usual care. Heterogeneity among included studies was assessed using the I^2^ statistic. We contacted authors for missing primary and secondary endpoint data. The statistical analyses were performed using the Cochrane Collaboration’s Review Manager Software (RevMan V.5.2.0).

## RESULTS

Of 4084 initial studies, 3,823 titles and abstracts were excluded, the main reasons for exclusion were, smoking cessation not delivered in a lung screening setting or population (wrong setting and wrong population), or no comparator group. This left 258 studies included in the full text screen, of these 13 studies were included (Figure 1). Reasons for exclusion were, no comparisons of cessation intervention, observational studies with no control group, abstracts, no smoking abstinence measures used as outcomes, review papers, study protocols and students’ dissertations. We also searched clinicaltrials.gov and the ISRCTN registry for ongoing trials in this area. We found 12 investigational trials comparing different smoking cessation interventions to control groups within a lung screening setting (20–31) (Table E6 Online Supplement).

### Overview of included studies

Ten studies were RCT (32–41) and 3 used an observational study with control group design (33, 42, 43) (Table 1). The studies were published between 2004 and 2022, involved a total of 5076 people who smoked, with sample sizes ranging from 18 to 1248. Median participant age ranged from 57 to 63 years, 61% of participants were male, mean number of cigarettes smoked per day by the population was 15.6±3.1 cigarettes.

**Table 1.** Summary of included studies

| Study (Design) | Setting | Smoking Cessation intervention | Intervention duration | Follow up | Control | Sample size | Results of Smoking Cessation group compared to control |
| --- | --- | --- | --- | --- | --- | --- | --- |
| Bade et al., 2016 (Observational with control) (44) | Screened with LDCT for Lung Ca | Personal smoking cessation counselling session delivered on the day of screening | 1 day | 12 months | Routine screening clinical care | 1236 | Quit rate: Intervention: 14.6% (85/611) vs Control: 6.7% (41/625). |
| Buttery et al., 2022 (RCT) (32) | Population attending for screening | 6 sessions of smoking cessation counselling plus pharmacotherapy (F2F) | 6 weeks | 3 months | Signposting and very brief advice | 115 | Quit rate Intervention: 29.2% (14/48) vs Control: 11% (4/36). Quit attempts Intervention: 54.1% (26/48) vs Control 33% (12/36). |
| Clark et al., 2004 (RCT) (33) | Attending LDCT lung Ca screening | Internet self-help materials including links to different cessation websites | 1 day | 12 months | Written take home materials | 171 | Quit rate intervention:13% (8/85) vs Control 7% (4/86). Quit attempts Intervention: 38% (23/85) vs Control 34% (20/86). |
| Ferketich et al., 2012 (RCT) (34) | Patients eligible for LDCT | Before CT(BCT), 1 F2F with medic, 12 sessions of tobacco dependence with NRT and weekly calls. | 12 weeks | 4 and 6 months | The same intervention but delivered after the CT scan (ACT) | 18 | Quit rates at 4 months BCT 33% (3/9) vs ACT 22.2% (2/9).<br>Quit rate at 6 months: BCT 22.2% (2/9) vs ACT 11.1% (1/9).<br>Quit attempts at 4 months: BCT 88.9% (8/9) vs ACT 100% (9/9).<br>Quit attempts at 6 months: BCT 66.7% (6/9) vs ACT 77.8% 7/8). |
| Lucchiari et al., 2020 (RCT) (35) | Sub population of | Two Armed RCT Arm 1- E-cigarettes with nicotine | 12 weeks | 6 months | Arm 3: Behavioural counselling | 155 | Quit rates: Arm 1: 16% (13/52), Arm 2: 19% (11/51), Control: 10% (7/52). |
|  | COSMOSII trial, large screening trial in Italy | Arm 2- Placebo (E-cigarettes without nicotine) |  |  | at weeks 1, 4, 8 and 12. |  | Exhaled CO: Arm 1: 12.01 ± 8.1ppm, Arm 2: 15.28 ± 11.4ppm, Control: 16.52 ± 10.2ppm<br>Reductions in cigarettes smoked per day Arm 1: 8.17± 6.5, Arm 2: 5.68 ± 7.9, Control: 5.82 ± 6.4. |
| Marshall et al., 2016 (RCT) (36) | Population of the Queensland screening trial | One session of behavioural support after screening, plus audio take home materials. | Missing | 12 months | Standard written materials with Quitline | 55 | Quit rate: Intervention: 14.3% (4/28) vs control: 18.5% (5/27)<br>Exhaled CO: Intervention 3.5% (1/28) vs control (11%) (3/27) |
| Pistelli et al., 2020 (Observational with control) (43) | Population of the ITALIA lung screening trial (Pisa Centre) | Six clinic visits within 3-5 months including counselling and pharmacotherapy and follow up at 6 and 12 months. | 5 months | 4 years | Normal screening clinical care | 425 | Higher odds of quitting in those who choose to attend the smoking cessation support (OR:3.16 (95% CI 1.63, 6.12). |
| Taylor et al., 2017 (RCT) (37) | Population undergoing LDCT screening at Georgetown University Medical Centre | Six sessions of weekly behavioural support telephone calls. | 6 weeks | 3 months | Information booklet, website, and smartphone app links. | 92 | Self-reported quit rate: intervention: 21.7% (10/46) vs Control 19.6% (9/46)<br>CO verified quit rates: Intervention: 17.4% (8/46) vs Control: 4.3% (2/46). |
| Taylor et al., 2022 (RCT) (38) | Population undergoing LDCT screening at Georgetown n University Medical Centre | Eight sessions of weekly behavioural support calls, plus 8-week supply of NRT patches. | 8 weeks | 3 months | Three behavioural support calls, plus 2-week supply of NRT patches. | 818 | Self-reported quit rates: Intervention 14.3% (58/409) vs control 9.7% (32/409).<br>CO verified quits: Intervention: 9.1% (37/409) vs Control 3.9% (19/409). |
| Tremblay et al., 2019 (RCT) (39) | LDCT Alberta screening trial. | Seven, weekly behavioural support telephone sessions. | 7 weeks | 12 months | Written information materials | 345 | Self-reported quit rates: Intervention: 14% (24/171) vs Control: 12.9% (22/174), CO verified quit rates: Intervention: 12.9% (22/171) vs Control: 10.9% (19/174) |
| van der Alast et al., 2011 (RCT) (40) | NELSON LDCT screening trial. | Computer tailored behavioural support messages | Missing | 24 months | Written information materials | 1248 | Self-reported quit rates Intervention: 12.5% (85/642) vs Control 15.6% (105/642). |
| Williams et al., 2022 (RCT) (41) | Population of HLP. Clinic visit and or LDCT | Six sessions of smoking cessation counselling plus pharmacotherapy, delivered via the telephone. | 6 weeks | 3 months | Signposting to local cessation services and very brief advice to quit. | 315 | Quit rates: Intervention 21.1% (32/152) vs control 8.9% (14/163).<br>Quit attempts: Intervention: 37.5% (57/152) vs Control: 22% (36/163). |
| Zeliadt et al., 2017 (Observational) (42) | Smokers offered LDCT screening | Two behavioural support calls, plus Quitline | 2 weeks | 1 month | Written information materials, plus Quitline | 83 | Quit rate: Intervention: 19% (5/27) vs Control: 7% (4/56)<br>Quit attempts: Intervention: 52% (14/27) vs Control: 45% (26/56) |

Studies investigated effectiveness of smoking cessation interventions as follows: single, in clinic session of behavioural counselling (44), single, in clinic session of behavioural counselling and audio materials (36), single in clinic behavioural counselling, plus NRT and subsequent weekly telephone calls (34), internet self-help materials (33), tailored information leaflet (40), 2 behavioural support calls and Quitline (42), multiple weekly behavioural support calls (37, 39), multiple weekly behavioural support calls and pharmacotherapy, (38, 41), multiple weekly behavioural support, in clinic plus pharmacotherapy (32, 43) or e-cigarettes (35). Timing of the intervention delivery varied across included studies from immediately on the day of the screening clinic to 5 months after screening, average length of interventions was 13.2±13.6 weeks.

A variety of comparisons were used for usual care (UC) groups e.g., signposting to local cessation clinics, written information leaflets and standard behavioural counselling, in addition to the intervention delivered before the lung health screening appointment. Of note Lucchiari et al., (35) used 4 sessions of behavioural counselling as their UC intervention, which is considerably more input compared to most UC interventions included.

All included studies used self-reported point prevalence smoking abstinence, either 30-day or 7-day, with 5 studies (34, 36–39) using exhaled carbon monoxide monitoring. Studies reported cessation outcomes at 1 month(42), 3 months (n=4)(32, 37, 38, 41), 4 and 6 months(34), 6 months(35), 12 months (n=4) (33, 36, 39, 44), 24 months (40), and 1 at 4 years (43). Five studies reported data on quit attempts(32-34, 41, 42) and 1 study reported data on reductions in daily cigarettes smoked (35).

### Description of included studies

Lucchiari et al., (35) conducted a 3 arm RCT, comparing the effect of behavioural counselling plus nicotine containing e-cigarettes, behavioural counselling plus nicotine free e-cigarettes (placebo) and behavioural counselling only (UC) on smoking behaviour in 155 smokers attending screening in Italy. Nine of the included studies were two-armed RCTs. Buttery et al., (32) randomised 115 smokers attending a targeted lung health check to immediate support including 6 sessions of behavioural counselling plus pharmacotherapy or very brief advice (VBA) to quit and signposting to local cessation services. Ferketich et al., (34), used a similar intervention (12 week tobacco cessation support, including pharmacotherapy), though this started a week after attending screening, rather than immediately. However, in this study the control group received the same smoking intervention delivered before the CT screening appointment (12 weeks of smoking cessation then attended screening on week 12) so it was not suitable for comparisons against “usual care”. Taylor et al., (37) randomised 92 smokers attending screening to receive 6 sessions of telephone behavioural counselling after participants had attended screening and received results, or a written information booklet (UC). A subsequent paper from that group randomised 818 smokers to receive intensive (8 telephone counselling sessions plus 8 weeks of NRT patch) or less intensive (3 counselling sessions, plus 2 weeks NRT patch), delivered 13 days post LDCT scan (38). Williams et al., (41) randomised 315 smokers attending a TLHC to receive 6 sessions of telephone behavioural support starting immediately, with pharmacotherapy prescriptions within 48 hours of participating in the TLHC, or VBA to quit and signposting to local services (UC). A telephone intervention was also utilised by Tremblay et al., (39) who randomised 354 smokers attending LDCT screening to 7 behavioural support calls delivered after participants had received screening results, or to written information leaflets only (UC). Three RCT used low-intensity interventions; Clarke et al., 2004 (33) randomised 171 smokers attending screening to receive internet materials or standard written materials delivered on the day of screening (UC). Marshall et al., 2016 (36) randomised 55 smokers attending screening to receive 1 session of behavioural support on the day of screening with audio take home materials, or written information plus Quitline (UC). Van der Alast et al., 2012 (40) randomised 1248 male smokers who were in the screening arm of the larger NELSON RCT, to receive computer tailored information leaflet or standard information leaflet after attendance to screening (UC).

The remaining 3 included studies used an observational design with a control group. Bade et al., 2016 (44), compared 12 month quit rates among 611 smokers attending screening who accepted the offer of one behavioural support session delivered before attendance to screening, to 625 smokers who received standard screening care. Pistelli et al., 2020 (43), explored quit rate among smokers at the Pisa centre in the ITALIA lung screening trial, screened participants were offered 6 smoking cessation clinic visits within 3-5 months of screening including counselling and pharmacotherapy (n=119), those who did not accept the offer of cessation went onto routine clinical care (n=306). Lastly, Zeliadt et al., 2017, used a waitlist design and gave smokers who recently attended a LDCT scan but had not received their results (n=27), two telephone behavioural counselling sessions, plus Quitline and compared quit rates to recent attendees to LDCT who had received their results (n=56).

### Effect of stop smoking interventions on quit rate compared to usual care

To analyse the effectiveness of smoking cessation interventions, we performed a meta-analysis on included RCT which compared quit rate between smoking cessation and usual care groups delivered in a screening context. Meta-analysis of 8 RCTs eligible for inclusion (32, 33, 35–39, 41) (n= 1,984) demonstrated that providing a smoking cessation intervention improved quit rates among current smokers enrolled in screening programmes compared with usual care (OR: 2.01, 95%: 1.49-2.72 p<0.001), Figure 2. We included data from Arm 1 (e-cigarette with nicotine) and Arm 3 (usual care) from the Lucchiari et al., (35) study, as this best answered the research question. Measures to assess quit rate varied between included studies, 3 studies used exhaled CO to measure quits (37–39), the remaining 5 studies used self-reported point prevalence smoking abstinence to assess quit rate (32, 33, 35, 36, 41).Data from two RCT were excluded from the meta-analysis, Ferketich et al.,(34) as they did not use a usual care group, rather explored the effect of timing of the same intervention and van der Aalst et al., (40) as their intervention group was too similar to their UC group (two types of information leaflet).

**Figure 2.**
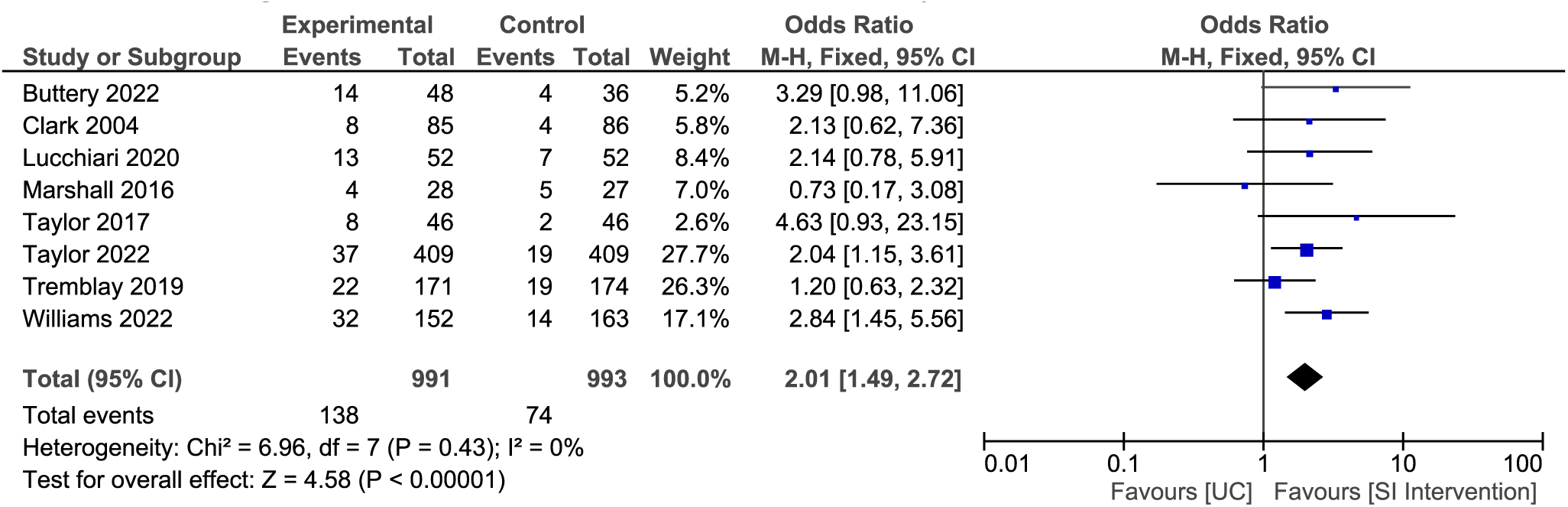
Effect of smoking cessation intervention vs usual care on quit rates Forest plot comparing quit rate among participants randomised to SC intervention compared with UC. Quit rates measured via self-reported (30-day or 7-day pp) or CO verification at follow up time periods ranging from 3 months to 12 months post intervention. Data from Arm 1 (E-cigarette with nicotine) and Arm 3 (UC) from Lucchiari et al’s 3 armed RCT was included in this analysis UC- Usual Care SI- Smoking Cessation

### Effect of intensive cessation interventions on quit rates

Meta-analysis of the 6 RCT (n=1,758) (32, 35, 37–39) that provided an intensive smoking cessation intervention, defined as ≥3 sessions with a smoking cessation advisor, nurse, or councillor+/- pharmacotherapies, demonstrated that intensive smoking cessation interventions yield greater quit rates compared to UC (OR: 2.11, 95% CI 1.53-2.90, p<0.001), Figure 3. We also conducted a meta-analysis among studies that provided pharmacotherapy (NRT, varenicline, e-Cigarettes), alongside behavioural counselling. Of note, only studies that provided intensive support were also the studies that provided pharmacotherapy. A meta-analysis of 4 RCTs(32, 35, 38, 41) (n=1,321), demonstrated that intensive smoking cessation support plus pharmacotherapy, yields higher quit rates compared to control among a screening population (OR: 2.40 95% CI 1.64-3.51, p <0.001) (Figure 4).

**Figure 3.**
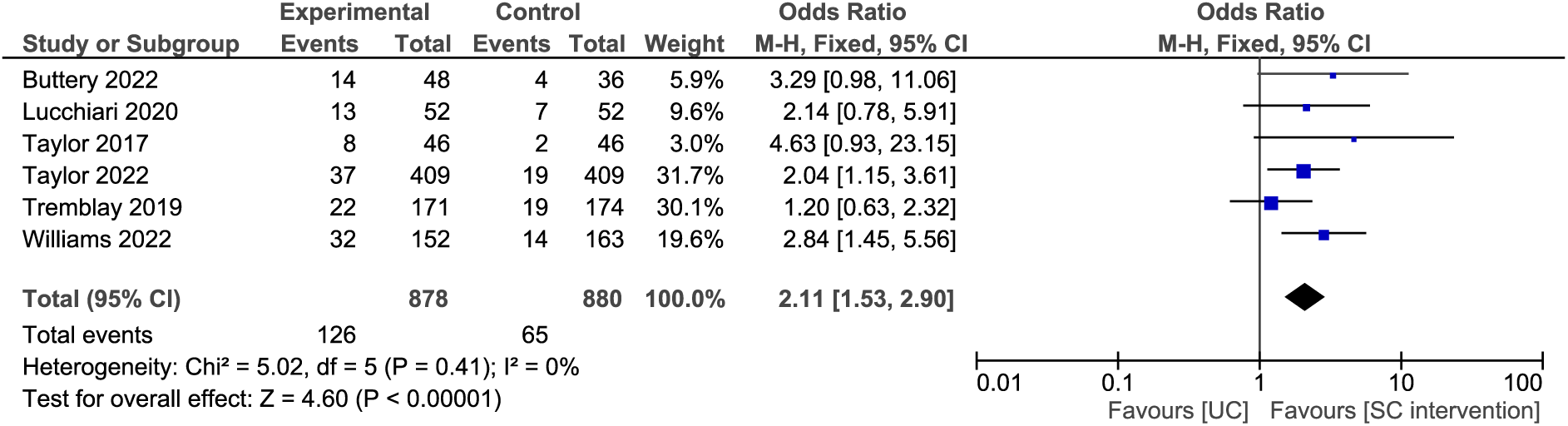
Effect of intensive smoking cessation intervention vs usual care on quit rates Forest plot comparing quit rates in studies that randomised participants to intensive SC interventions (≥3 behavioural counselling sessions compared with UC. Quit rates measured via self-reported (30-day or 7-day pp) or CO verification at follow up time periods ranging from 3 months to 12 months post intervention. Data from Arm 1 (E- cigarette with nicotine) and Arm 3 (UC) from Lucchiari et al’s 3 armed RCT was included in this analysis. UC- Usual Care SI- Smoking Cessation

**Figure 4.**
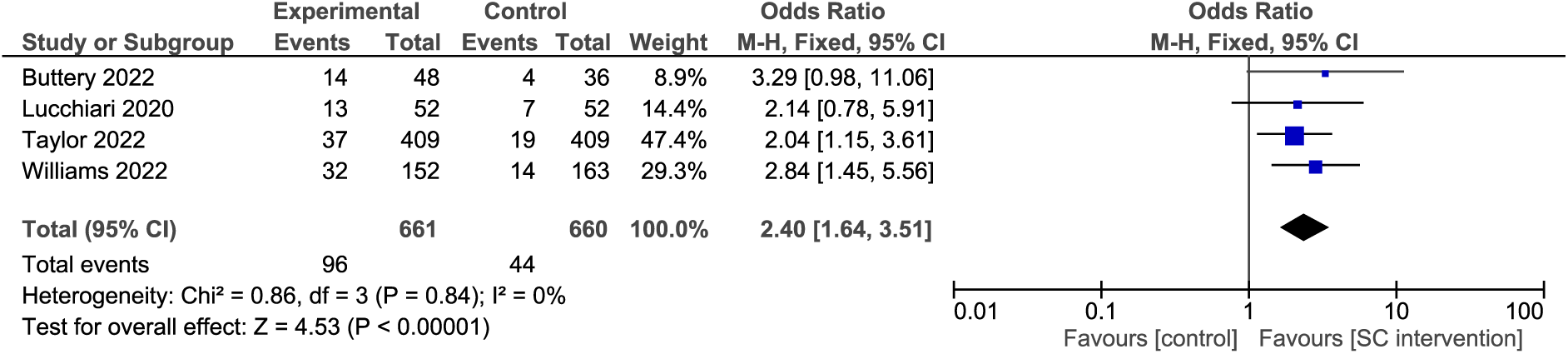
Effect of intensive smoking cessation with pharmacotherapy vs usual care on quit rates Forest plot comparing quit rates in studies that randomised participants to intensive SC interventions (≥3 behavioural counselling sessions and pharmacotherapy compared with UC. Quit rates measured via self-reported (30- day or 7-day pp) or CO verification at follow up time periods ranging from 3 months to 6 months post intervention. Data from Arm 1 (E-cigarette with nicotine) and Arm 3 (UC) from Lucchiari et al’s 3 armed RCT was included in this analysis UC- Usual Care SI- Smoking Cessation

### Effect of intensive interventions compared to non-intensive interventions on quit rate

A meta-analysis of 2 RCT (35, 38), (n=922) investigating the effect of intensive interventions compared to usual care, in the case of these two studies their UC interventions would be classed at a non-intensive smoking cessation intervention itself. Results from the analysis demonstrated that intensive smoking cessation interventions compared to non-intensive smoking cessation interventions, results in higher quit rates (OR: 2.07, 95%CI 1.26-3.40 p=0.004), Figure 5.

**Figure 5.**
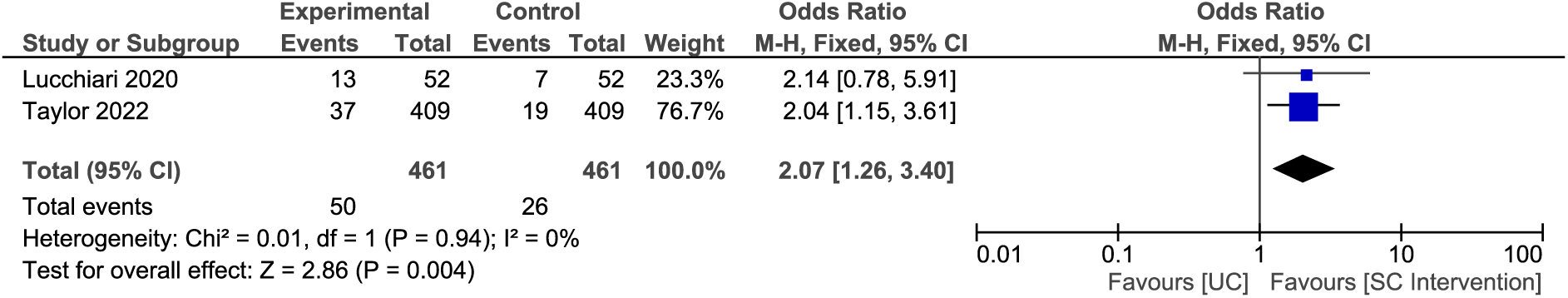
Effect of intensive vs non-intensive smoking cessation interventions on quit rates Forest plot of self-reported quit rates in two studies that randomised participants intensive SC interventions compared with non- intensive SC interventions Quit rates measured via self-reported pp at 6 month follow up, and CO verification at 3 month follow up. Data from Arm 1 (E-cigarette with nicotine) and Arm 3 (UC) from Lucchiari et al’s 3 armed RCT was included in this analysis UC- Usual Care SI- Smoking Cessation

### Effect of non-intensive interventions on quit rate

A meta-analysis of 2 RCT (33, 36) (n= 226) investigating non- intensive smoking cessation interventions (≤2 behavioural counselling sessions) did not find higher quit rates compared to UC, (OR: 0.90, 95% CI 0.39-2.08 p=0.80), Figure 6.

**Figure 6.**
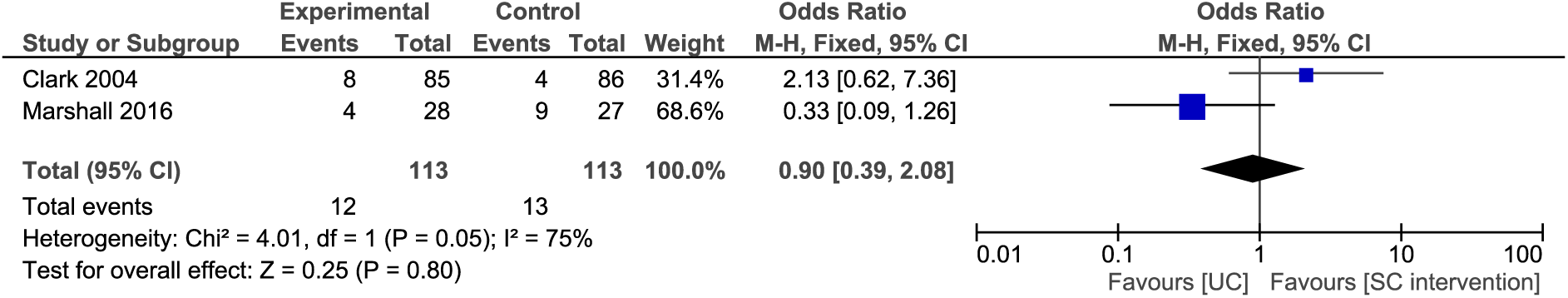
Effect of non-intensive smoking cessation intervention vs usual care on quit rates Forest plot of self-reported quit rates in two studies that randomised participants to non- intensive SC interventions compared with UC (≤2 counselling sessions, online materials, audio self-help materials). Quit rates measured via self-reported pp at 12 month follow up. UC- Usual Care SI- Smoking Cessation

### Effect of interventions on quit attempts

The impact of smoking cessation interventions on the number of participants reporting quit attempts could be compared in 3 RCTs. (32, 33, 41)(n=570), showing that smoking cessation interventions produce significantly higher quit attempts compared to UC interventions, within a screening context (OR: 1.85, 95%CI 1.28-2.66, p<0.001) (Figure E1. Online Supplement).

### Risk of bias and evidence quality

Risk of bias (ROB) and evidence quality was judged using the Cochrane risk-of-bias assessment tool (45). There was large variation in the risk of bias among included studies, with 4 studies assessed as having low risk of bias (32, 35, 38, 41). We assessed 4 studies as having an unclear risk of bias (33, 37, 39, 40), mainly attributed to unclear methods of blinding of outcome assessors and unclear methods of randomisation. The remaining 5 studies were deemed high risk of bias (34, 36, 42–44), attributed to the observational study design in three of the studies (42–44), with no attempt to blind outcome assessors, poor allocation concealment, selective reporting and low sample sizes seen in two of the studies (36, 42)(Figure 7). A more detailed description of risk of bias domains and scoring can be found on the online supplement (Table E1, online supplement).

**Figure 7.**
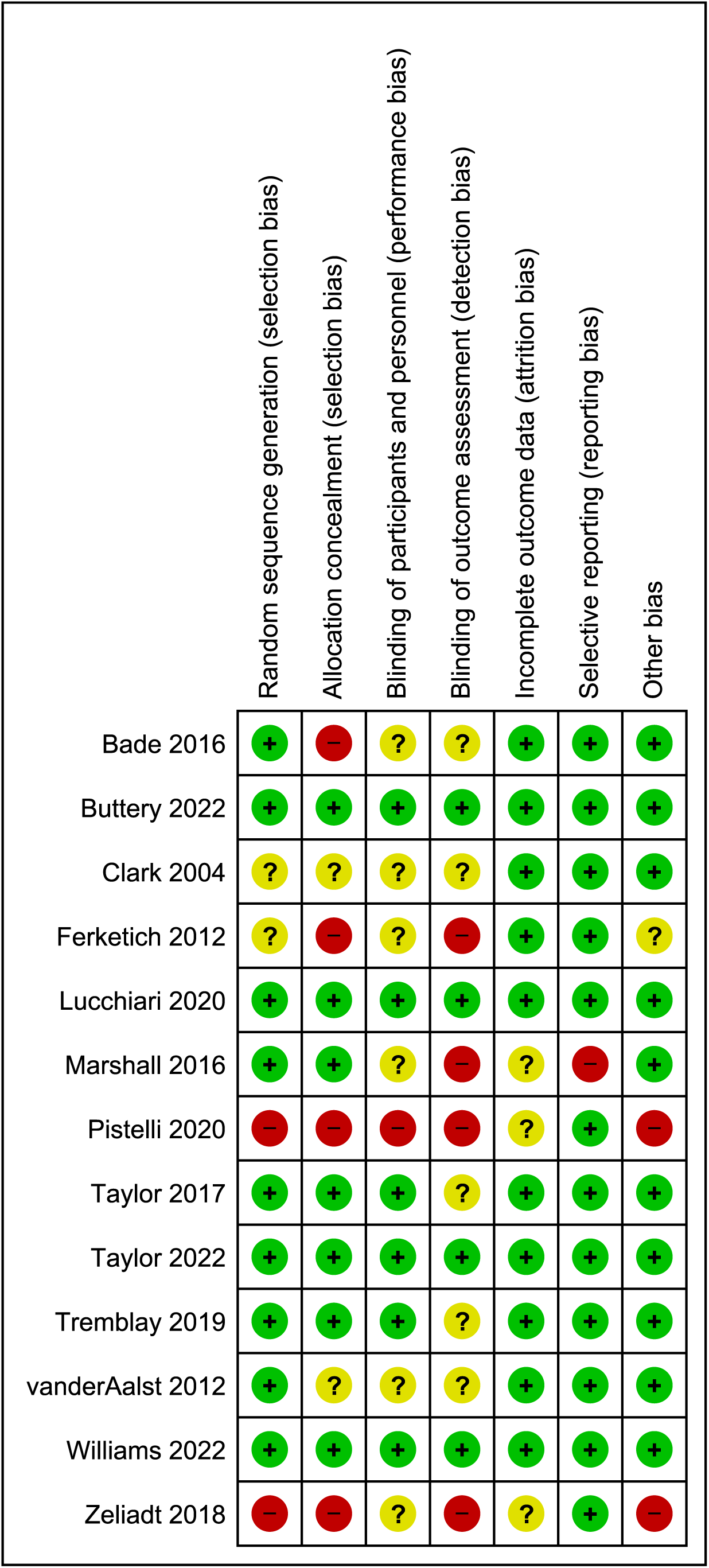
Risk of bias of assessment of included studies (45).

Using the GRADE criteria (18), we found overall moderate quality evidence to support inclusion of smoking cessation interventions into lung cancer screening, attributed to strong association of results, directness of the evidence in terms of assessing our primary endpoint (quit attempts) and the precision of results (large sample size of included participants) (Table E2, online supplement). For intensive smoking cessation interventions, the GRADE criteria found high quality evidence supporting embedding intensive smoking cessation interventions into lung screening, compared both to usual care and to low intensity interventions, gain attributed to strong association of results, directness of the evidence in terms of assessing primary endpoint (quit attempts), the precision of results (large sample size) and moderate inconsistency (heterogeneity) (Table E4, online supplement). In terms of non-intensive smoking cessation interventions, the GRADE criteria found low certainty evidence for non-intensive interventions, attributed to low and unclear risk of bias, large heterogeneity across included studies and the low sample sizes of Marshall et al., 2016 (36) and Zeliadt et al., 2018 (42) (Table E5, online supplement).

## DISCUSSION

This systematic review addressed the effectiveness of smoking cessation intervention strategies delivered as a component of lung health screening programmes. Moderate quality evidence supports the embedding of smoking cessation interventions within screening programmes. Intensive interventions and those including pharmacotherapy support (NRT, varenicline or E-cigarettes) were supported by high quality evidence and appeared to be more effective at increasing quit rates. Where smoking cessation intervention intensity was low, the evidence supporting the recommendation of these strategies was low, suggesting a minimum intervention threshold needed to have an effect.

Given the well-established health and social harms from smoking, the value of lung cancer screening programmes will be enhanced by embedding smoking cessation interventions into them. Modelling suggests that providing behavioural cessation advice, plus pharmacotherapy can reduce the cost per quality adjusted life year (QALY) of a lung screening programme by up to 50%(13). The economic evaluation of the Taylor et al., 2022 (38) RCT comparing 8-week intensive intervention to 2-week minimal intervention, also estimated greater lifetime cost savings with cessation support. The incremental costs per QALY gained were estimated to be larger with the 8-week intensive counselling (46). Results from our current review, suggest that providing more support (multiple counselling sessions and pharmacotherapy) compared with less-intensive approaches, may be more favourable for a screening population. However, there were only limited direct comparison data, leading to only a recommendation of moderate certainty. Two RCT found increases in quit rates with more intensive smoking cessation support compared with less intensive support. Lucchari et al., 2020 found that nicotine (e-cigarettes) with 4 sessions of behavioural counselling was more effective than behavioural support alone(35). Taylor 2020 et al., demonstrated self-reported and CO verified quit rates were significantly higher among smokers who received 8 counselling sessions plus 8 weeks of NRT patch vs 3 sessions plus 2-week supply of NRT (38), which further supports the argument that smoking cessation interventions delivered in a screening context should provide multiple sessions and offer pharmacotherapy provision. We also note that all the intensive interventions included pharmacotherapy, so it is not possible from the available data to disentangle these two effects, although best practice is anyway for pharmacotherapy to be offered with behavioural support.

E-cigarettes have been supported as a stop smoking aid by a recent UK NIHR Health technology assessment(47) and a living Cochrane review finds high certainty evidence that nicotine containing e-cigarette use increases quit rates compared to NRT(48). However, this may in part be due to the typically longer duration of use of e-cigarettes compared to medicinal forms of NRT and other pharmacological agents. This probably reduces the risk of relapse to smoking, but longer-term use of e-cigarettes is itself likely to cause some health harms. Individuals should be advised to quit vaping in due course if they can, though not at the expense of going back to smoking.

To date there has been one previous systematic review aimed at investigating effective smoking cessation interventions delivered during LDCT screening. Iaccarino et al., (49), conducted their review in 2019, concluding that the data was insufficient to recommend one smoking cessation intervention over another. Our current review also contains statistical pooled analyses of included studies enabling us to draw firmer conclusions from the data.

This review has certain limitations. The primary endpoint of smoking cessation was assessed at a range of time points in the studies included. The immediacy of intervention also varied as well as what was considered to be usual care(35) . This led to a high level of heterogeneity between the studies included in the meta-analysis, however this is to an extent addressed in the different analyses presented, comparing both any additional smoking cessation intervention vs usual care and more intense intervention vs less. Despite having no search limitations based on location, all included studies were conducted in high income countries, so generalisability to middle and low-income countries need to be considered. Additionally, tobacco use habits are changing and will be influenced by the social and cultural factors. As the studies included in this analysis were all conducted in Europe/USA, the extent to which these findings can be applied globally is unclear. Although not included in the meta-analysis, the findings of observational studies also support the conclusion that smoking cessation is effective in this context.

This review demonstrates that providing smoking cessation within lung health screening programmes increases quit rates. Given that smoking cessation is arguably the most successful and cost-effective intervention for reducing tobacco induced diseases, it is vital that smokers who attend screening clinics are offered support. This not only reduces risk for the individual but potentially increases the cost effectiveness of the screening intervention (13, 46). The population who attend lung screening clinics will typically have a high degree of nicotine dependence and addiction, in addition to a higher risk of not only frequent respiratory exacerbations and ill health(50), but also social isolation and loneliness(51, 52). Because of that, we suggest that offering pharmacotherapy alongside behavioural counselling support, should be mandated when designing pathways to support smoking cessation within lung cancer screening. Finally, it is apparent that more research is needed within this area, particularly aimed at intensiveness and timing of the interventions.

## CONCLUSION

Lung health screening settings are a key environment for delivering smoking cessation. All screening programmes should provide evidence-based smoking cessation alongside screening, and efforts should be made to provide sustained counselling support and provide pharmacotherapy.

## AUTHOR CONTRIBUTION

PJW, KEJP, SMA, NSH and AMP conducted screening and data extraction, PJW and AAL conducted ROB analysis, PJW conducted pooled analyses with guidance from NSH and AAL. PJW produced the first draft to which all authors contributed. All authors have reviewed and approved the final manuscript and NSH is the guarantor.

## CONFLICTS OF INTEREST

PW, KEJP, SMA, AMP, SB & MP have no conflicts of interest to declare, NSH is medical director of Asthma+Lung UK and chair of Action on Smoking and Health, AAL is a trustee of Action on Smoking and Health.

## FUNDING

This work was supported by RM Partners, West London Cancer Alliance, hosted by The Royal Marsden NHS Foundation Trust and Royal Brompton and Harefield Charities. KEJP was supported by the Imperial College Clinician Investigator Scholarship. The funders had no input into data analysis or the writing of this manuscript.

## TRANSPARENCY DECLARATION

Nicholas Hopkinson, the manuscript’s guarantor affirms that the manuscript is an honest, accurate, and transparent account of the study being reported; that no important aspects of the study have been omitted; and that any discrepancies from the study as planned (and, if relevant, registered) have been explained.

## Supporting information

the precision of results (large sample size) and moderate inconsistency (heterogeneity) (Table E4, online supplement

## Data Availability

All data produced in the present study are available upon reasonable request to the authors

## Notes

### Clinical Protocols

https://www.crd.york.ac.uk/prospero/display_record.php?RecordID=242431

### Funding Statement

This study did not receive any funding

