## Supplementary material for "Strategies to deliver smoking cessation interventions during targeted lung health screening - a systematic review and meta-analysis": the precision of results (large sample size) and moderate inconsistency (heterogeneity) (Table E4, online supplement

Parris J Williams ^1,3^, Keir EJ Philip ^1^, Saeed M Alghamdi ^4^, Alexis M Perkins ^1^, Sara C Buttery ^1^, Michael I Polkey^1^, Anthony A Laverty ^2^, Nicholas S Hopkinson^1^

^1^National Heart and Lung Institute, Imperial College

^2^Public Health Policy Evaluation Unit, School of Public Health, Imperial College London, London, UK

^3^Royal Brompton and Harefield Hospitals

^4^ Clinical technology Department, Respiratory Care Program, College of Applied Medical Science, Umm-Al Qura University.

Page 1: Figure E1. Forest plot comparing quit attempts between smoking cessation intervention arms and usual care arms.

Pages 2-4: Table E1, Risk of bias domains

Pages 5-6 Tables E2-E5, Grade Assessments

Pages 6-14 Table E6, Summary of ongoing clinical trials

**Forest Plot comparing quit attempts between smoking cessation intervention arms and usual care arms**

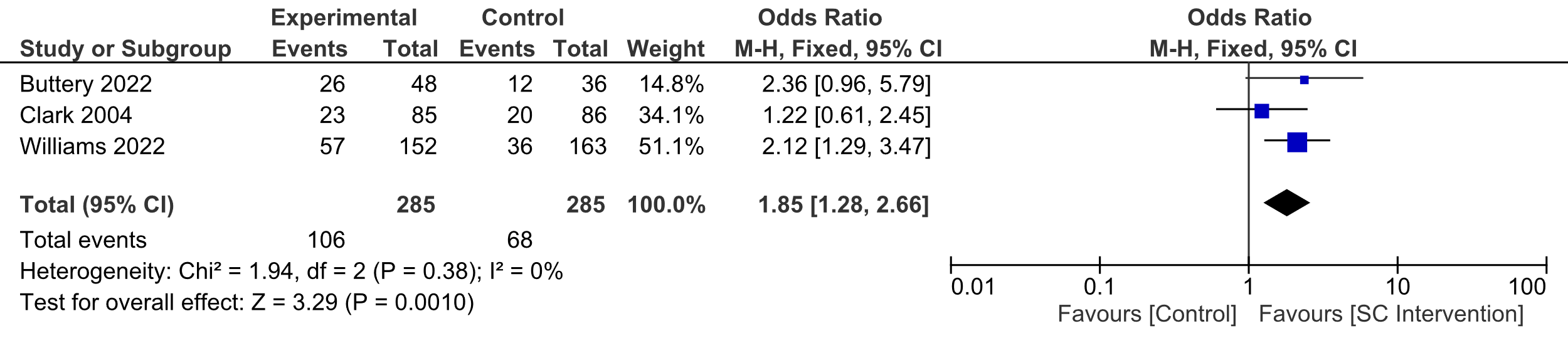

**Figure E1. Forest plot comparing quit attempts between smoking cessation interventions versus control. Quit attempts measured by self-reported data, follow up time periods ranging from 3- 12 months post intervention.**

**UC- Usual Care**

**SI- Smoking Cessation**

**Table E1. Risk of bias domains**

|  | Random Sequence generation | Allocation Concealment | Blinding- Participants & Personnel | Blinding of outcome assessors | Incomplete outcome data | Selective reporting | Other | Overall Score |
| --- | --- | --- | --- | --- | --- | --- | --- | --- |
| Bade et al., 2016 (Observational with control) (32) | Low: Use of electronic randomisation system in 2012 study | High: Participants opted into the intervention group | Unclear: Not stated if study personnel were blinded to outcome | Unclear not clear if outcome or assessors were blinded. | Low: Use of multiple imputations for missing data | Low: Both primary and secondary outcomes reported | Low: None | High |
| Buttery et al., 2022, (RCT) (20) | Low: Use of sealed envelope, randomised by day of the week | Low: Only nurses delivering intervention, participants and nurses delivering TLHC aware of group allocation. | Low: Study personnel conducting follow up unaware of group allocation. | Low: All outcome assessors blinded. | Low: use of ITT, those who were lost to follow up recorded as current smokers | Low: Both primary and secondary outcomes reported | Low: None | Low |
| Clark et al., 2004, (RCT) (21) | Unclear: Not stated how randomisation occurred. | Unclear: Not stated how allocation concealment occurred. | Unclear: Not stated if blinding of personnel occurred. | Unclear: not stated if outcome assessors/ analysists were blinded | Low: use of ITT, those who were lost to follow up recorded as current smokers | Low: Both primary and secondary outcomes reported | Low: None | Unclear |
| Ferketich et al., 2012, (RCT), (22) | Unclear: stated randomisation occurred but not how was conducted | High: No allocation concealment occurred | Unclear: blinding of personnel occurred. | High: No blinding of outcome assessors/ analysists | Low: No missing data (small sample size) | Low: Both primary and secondary outcomes reported | Unclear Small sample size but stated as a pilot study. | High |
| Lucchiari et al., 2020, (RCT) (23) | Low: Permuted block design 40 blocks of 6 subjects. | Low: Use of placebo group | Low: Participants in Arms 1,2 blinded alongside study personnel. | Low: Stated as double blind | Low: use of ITT, those who were lost to follow up recorded as current smokers | Low: Both primary and secondary outcomes reported | Low: None | Low |
| Marshall et al., 2016, (RCT) (24) | Low: 1:1 randomisation method | Low: Group allocation concealment | Unclear: not stated if personnel blinded | High: No blinding of outcome assessors/ analysists | Unclear: not stated if any data was missing | High: They did not report CO verified quit data | Low: None | High |
| Pistelli et al., 2020, (observational with control) (31) | High: No sequence generation occurred | High: No allocation concealment occurred | High: No blinding occurred | High: No blinding occurred | Unclear: Use of ITT but not stated how missing data was handled. | Low: Both primary and secondary outcomes reported | High: Potential risk of social desirability bias | High |
| Taylor et al., 2017, (RCT) (25) | Low: Permuted block design, blocks of 4 stratified by site. | Low: use of computer allocator | Low: Study personnel blinded | Unclear: not stated if outcome assessors/ analysists were blinded | Low: Use of ITT, those who were lost to follow up recorded as current smokers | Low: Both primary and secondary outcomes reported | Low: None | Unclear |
| Taylor et al., 2022, (RCT) (26) | Low: Permuted block design, blocks of 4 stratified by site. | Low: use of computer allocator | Low: Study personnel blinded | Low: outcome assessors blinded | Low: Use of ITT, those who were lost to follow up recorded as current smokers | Low: Both primary and secondary outcomes reported | Low: None | Low |
| Tremblay et al., 2019, (RCT) (27) | Low: 1:1 stratified by CT result and intention to quit | Low: use of computer allocator | Low: Study personnel blinded | Unclear: not stated if outcome assessors/ analysists were blinded | Low: Use of ITT, those who were lost to follow up recorded as current smokers | Low: Both primary and secondary outcomes reported | Low: None | Unclear |
| van der Alast et al,. 2012, (RCT) (28) | Low: 1:1 randomisation method | Unclear: Not stated how allocation concealment occurred. | Unclear: not stated if personnel blinded | Unclear: not stated if outcome assessors/ analysists were blinded | Low: Use of ITT, those who were lost to follow up recorded as current smokers | Low: Both primary and secondary outcomes reported | Low: None | Unclear |
| Williams et al., 2022, (RCT), (29) | Low: Use of sealed envelope, randomised by day of the week | Low: Only nurses delivering intervention, participants and nurses delivering TLHC aware of group allocation. | Low: Study personnel conducting follow up unaware of group allocation. | Low: All outcome assessors blinded. | Low: use of ITT, those who were lost to follow up recorded as current smokers | Low: Both primary and secondary outcomes reported | Low: None | Low |
| Zeliait et al., 2017, (Observational with control) (30) | High: No sequence generation occurred | High: No allocation concealment occurred | Unclear: not stated if personnel blinded | High: No blinding out outcome assessors | Unclear: not stated how missing data was handled | Low: Both primary and secondary outcomes reported | High: probable recruitment bias | High |

Table E2. Grade assessment: Smoking cessation interventions vs control for self-reported quit rates.

| Participants (Studies) | Risk of Bias | Inconsistency | Indirectness | Imprecision | Other Considerations | Overall Certainty |
| --- | --- | --- | --- | --- | --- | --- |
| 5,079 (10 RCT’s & 3 Observational) | Serious: Majority of studies had either unclear or high risk of bias | Serious: Heterogeneity between studies, variation in quit rates. | Not Serious: All studies directly answer the healthcare question | Not Serious: Large sample size, with large number of events | Minimal publication bias, strong association of results | Moderate |

Table E3. Grade Assessment: Smoking cessation interventions vs control for self-reported quit attempts

| Participants (Studies) | Risk of Bias | Inconsistency | Indirectness | Imprecision | Other Considerations | Overall Certainty |
| --- | --- | --- | --- | --- | --- | --- |
| 701 (4 RCT’s & 1 Observational) | Serious: Majority of studies had either unclear or high risk of bias | Serious: Heterogeneity between studies, variation in quit rates. | Not Serious: All studies directly answer the healthcare question | Not Serious: Large sample size, with large number of events | Some publication bias, Strong association of results. | Moderate |

Table E4. Grade Assessment: Intensive smoking cessation interventions vs control for self-reported quit rate

| Participants (Studies) | Risk of Bias | Inconsistency | Indirectness | Imprecision | Other Considerations | Overall Certainty |
| --- | --- | --- | --- | --- | --- | --- |
| 1,910 (7 RCT’s) | Serious: Some of the studies had either unclear or high risk of bias | Not Serious: low Heterogeneity between studies, majority of studies reported increases in quit rates. | Not Serious: All studies directly answer the healthcare question | Not Serious: Large sample size, with large number of events | Minimal publication, Strong association of results. | High |

Table E5. Grade Assessment: Non-intensive smoking cessation interventions vs control for self-reported quit rate

| Participants (Studies) | Risk of Bias | Inconsistency | Indirectness | Imprecision | Other Considerations | Overall Certainty |
| --- | --- | --- | --- | --- | --- | --- |
| 2,829 (3 RCT’s & 2 Observational) | Serious: Majority of the studies had either unclear or high risk of bias | Serious: large heterogeneity between studies, variations in quit rate success | Not Serious: All studies directly answer the healthcare question | Not Serious: Large sample size, with large number of events | Publication bias detected in 3 of the studies, weaker association of results. | Low |

Table E6. Ongoing Clinical Trials investigating smoking cessation interventions in the context of Lung Cancer Screening

| Trial | Primary Investigator | Intervention | Comparison | Primary Outcomes | Secondary Outcomes | Expected Completion date |
| --- | --- | --- | --- | --- | --- | --- |
| Program for lung cancer screening and tobacco cessation (PULTO)  Sequential, multiple assignment, randomized trial (SMART) design. NCT02597491 (USA).  https://clinicaltrials.gov/ct2/show/NCT02597491) | Joseph, A | Arm 2: Telephone tobacco longitudinal care (TLC)  Arm 3: TLC plus pharmacotherapy | Arm 1: 8 weeks of tobacco cessation | 6 months continued tobacco abstinence at 18 months post randomisation | Intensiveness of TLC on abstinence (monthly vs quarterly follow up)  How CT results moderate smoking outcomes. | 30/11/2022 (Results not published) |
| Capitalizing on a teachable moment to promote smoking cessation, RCT. NCT02276664 (USA). (<https://clinicaltrials.gov/ct2/show/study/NCT02276664>) | Brandon, T | Behavioural self-help booklet intervention (developed via focus groups) | Standard information booklet | 7-day point prevalence abstinence 9 months post randomisation |  | 21/06/2019 (Results not published) |
| Personalized Smoking Cessation Tool Based on Patient Lung CT Image, RCT. NCT03087617 (USA). (https://clinicaltrials.gov/ct2/show/NCT03087617) | McEvoy, C | Arm 1: Usual care + smoking cessation report that includes data analysis from a CT lung cancer screening exam.  Arm 2: Usual care plus behavioural counselling.  Arm 3: Usual care + smoking cessation report + behavioural counselling. | Usual care for lung cancer screening patients and Quitline | Reports impact on participants calling Quitline number (3 weeks are treatment). | Number of quit attempts (3 weeks after treatment) | 23/08/2021 (Results not published) |
| Yorkshire Enhanced Stop Smoking study (YESS), RCT. NCT03750110. (UK). (https://clinicaltrials.gov/ct2/show/NCT03750110) | Murray, R | Standard smoking cessation support in line with NICE guidelines plus personalised feedback from CT result. | Standard smoking cessation support inline with NICE guidelines | 7-day point prevalent CO validated smoking cessation (3 months post CT scan). | Continuous smoking abstinence at three months.  Smoking abstinence measures at 4 weeks and 12 months post CT.  Changes in perceived lung cancer risk, motivation to quit smoking, confidence, and efficacy beliefs of stopping smoking, Self -reported changes in health at 4 weeks, 3 months, and 12 months post CT. | 21/03/2022 (Results not published) |
| Implementing Tobacco Treatment in Low Dose CT Lung Cancer Screening Sites, RCT. NCT03315910 (USA). (https://clinicaltrials.gov/ct2/show/NCT03315910) | Ostroff, J | Arm 1: 2 motivational interviewing sessions (telephone or face to face)  Arm 2: Nicotine Lozenge, 6 cartons of 2 mg lozenges.  Arm 3: Nicotine patch, 6 weeks.  Arm 4: Message framing, participants receive a standardized message that emphasizes either the benefits of quitting (gain-framed) or the risks of continuing to smoke (loss-framed). | Enhanced standard care: education and follow-up cessation counselling during their first session within 1 month of randomization or during the shared decision-making discussion + self help materials and Quitline | Smoking abstinence (self-reported and Bio verified 6 months post randomisation. | Quit attempts and reductions in cigarettes per day. | October 2024 |
| Implementation of Smoking Cessation Within NCI Community Oncology Research Program (NCORP) Sites (OaSiS), RCT. NCT03291587 (USA) (https://clinicaltrials.gov/ct2/show/NCT03291587) | Foley, K | Tobacco coaching sessions, 6 sessions every 4-6 weeks over 8 months | Usual screening clinical care | 7-day sustained smoking abstinence 6 months after treatment | Bio verified smoking abstinence (6 months follow up)  Short term smoking abstinence ( 3 month follow up). | 25/07/2022 (Results not published) |
| Smoking Cessation Intervention During Low Dose CT (LDCT) Screening for Lung Cancer, RCT. NCT03059940 (USA). (https://clinicaltrials.gov/ct2/show/NCT03059940) | Cinciripini, P | Quitline + Rx (Arm1): LDCT provider and patient discuss options for pharmacotherapy Quitline referral. 5 smoking cessation counselling sessions over the next 12 weeks.  Integrated Care (IC) Group (Arm2): Participant referred to Tobacco Treatment Program (TTP). TTP provides 4-8 counselling sessions and pharmacotherapy over a 10–12-week period, | Quitline group: Referral to Quitline for counselling +NRT for at least 5 sessions | Smoking abstinence at 6 months |  | 01/06/2023 |
| Promoting Smoking Cessation in Lung Cancer Screening Through Proactive Treatment (PROACT), RCT NCT03612804 (USA). (https://clinicaltrials.gov/ct2/show/NCT03612804) | Zeliadt, S | Two sessions of proactive cessation care, including cessation medications and behavioural telephone counselling | Standard clinical screening care | Self-reported abstinence from smoking 12 months post randomisation |  | 30/04/2023 |
| CONNECT Smoking cessation and lung cancer screening, RCT, NCT04149249 (USA). (https://clinicaltrials.gov/ct2/show/NCT04149249) | Walsh, J | View and participate in the interactive Video Doctor about Smoking Cessation before LCS appointment | Standard information leaflet about smoking cessation. | 30-day abstinence, Self-reported quit attempts, percentage of participants who attend cessation clinics. | Change in participation rate | 31/05/2023 |
| Comparing smoking cessation interventions among underserved patients referred for lung cancer screening, RCT NCT04798664 (USA) (https://clinicaltrials.gov/ct2/show/NCT04798664) | Scott D Halpern | Arm 1: free access to nicotine replacement therapy (NRT) and/or reimbursement of up to $300 for any smoking cessation medications (varenicline/Chantix or bupropion/Zyban)  Arm 2: Arm 1 plus an incentive plan in which participants will be informed of their eligibility to earn $100, $200, and $300 if they submit negative tests for nicotine metabolites at 2 weeks, 3 month and 6 months following their quit date, respectively.  Arm 3: Participants receive all aspects of Arm 2 plus an intervention to promote episodic future thinking (EFT), Patients will practice using EFT cues to envision the "future is now" between the time of enrolment and the quit date, and will then receive cues from the quit date through the end of the intervention period, 6 months later. | Standard cessation care, Ask, Advise, Refer | Biochemically confirmed smoking abstinence sustained for 6 months | Self -report smoking abstinence and Bio confirmed at 2 weeks, 3 months, 6 months, 12 months | December 2024 |
| Tobacco treatment in the context of lung cancer screening, RCT NCT03927989, (USA). (https://clinicaltrials.gov/ct2/show/NCT03927989) | Rojewski, A | Advice to quit and brief discussion of tobacco use plus dual nicotine replacement therapy plus 8 weeks of gain-framed text messages tailored to lung screening | Brief advice to quit smoking prior to Lung screening | Self-reported 7 day PP smoking abstinence 8 weeks after treatment | End of Study Abstinence Rates  3 months follow up | August 2023 |
| Assessing the integration of Tobacco Cessation Treatment into Lung Cancer Screening (LCS), (ScreenASSIST), RCT (NCT03611881), (USA). https://clinicaltrials.gov/ct2/show/NCT03611881) | Park, E | Arm 1: 4 weeks of counselling + 2 weeks of NRT + local referral  Arm 2: 4 weeks of counselling + 8 weeks of NRT + local referral  Arm 3: 8 weeks of counselling + 2 weeks of NRT + local referral  Arm 4: 8 weeks of counselling + 8 weeks of NRT + local referral  Arm 5: 4 weeks of counselling + 2 weeks of NRT + no referral  Arm 6: 4 weeks of counselling + 8 weeks of NRT + no referral  Arm 7: weeks of counselling + 2 weeks of NRT + no referral  Arm 8: 8 weeks of counselling + 8 weeks of NRT + no referral | No control | Self-reported past 7-day smoking abstinence (6 months post treatment) | Reduction in cigarettes per day , intention to quit. | 01/02/2023 |
